# Memory B cell and humoral responses elicited by Sputnik V in naïve and COVID-19-recovered vaccine recipients

**DOI:** 10.1101/2021.10.13.21264894

**Authors:** Maria G Byazrova, Sergey V Kulemzin, Ekaterina A Astakhova, Tatyana N Belovezhets, Grigory A Efimov, Anton N Chikaev, Ilya O Kolotygin, Andrey A Gorchakov, Alexander V Taranin, Alexander V Filatov

## Abstract

The development of effective vaccines against SARS-CoV-2 remains a global health priority. Despite extensive use, the effects of Sputnik V on B cell immunity need to be explored in detail. We show that B memory cell (MBC) and antibody responses to Sputnik V were heavily dependent on whether the vaccinee had a history of SARS-CoV-2 infection or not. *In vitro* stimulated MBCs from previously infected recipients of Sputnik V secreted a significant amount of anti-RBD IgG both on days 28 and 85 from the beginning of vaccination. These antibodies demonstrated robust neutralization of the Wuhan Spike-pseudotyped lentivirus. In the naïve group of vaccinees, the level of anti-RBD IgG secretion was five- to six-fold reduced compared to that of the recovered group, and maximum virus neutralization (Wuhan spike) was achieved only on day 85. Sera from all the recovered and most naïve Sputnik V recipients were neutralizing against the ancestral Wuhan and mutant B.1.351 viruses. Thus, our in-depth analysis of MBC responses in Sputnik V vaccinees complements traditional serological approaches and may provide important outlook into future B cell responses upon re-encounter with the emerging variants of SARS-CoV-2.

## Introduction

Presently, the therapeutic options for COVID-19 patients remain limited, emphasizing the necessity of concerted mass vaccination campaigns to counteract the pandemic. An ideal vaccine must induce long-lasting protective cellular and humoral immunity, which should translate into reduced rates of infection and mortality. Importantly, an ideal vaccine should, in addition, retain activity against emerging viral lineages.

Three anti-SARS-CoV-2 vaccines, Moderna mRNA-1273, BioNTech BNT162b2, and Janssen Ad26.COV2.S, are now being extensively used around the world and have received the most public attention and validation (Goel et al., 2021), while less is known about immunity after Sputnik V vaccination (Logunov et al., 2021). Although humoral responses to Sputnik V have recently been reported for a limited number of study participants (Ikegame et al., 2021; Rossi et al., 2021), data regarding the B cell response in Sputnik V-vaccinated subjects are presently lacking. Clearly, these data are central to the comprehensive assessment of current vaccines (Goel et al., 2021), provide important clues to the development of new vaccines, and impact the epidemiological models of immunity.

In late 2020, multiple SARS-CoV-2 lineages were reported across the globe, of which B.1.1.7, B.1.351, B.1.1.248, and B.1.617 are now referred to as variants of concern (VOCs) (Davies et al., 2021; Tegally et al., 2021). B.1.351 and B.1.1.248 display profound resistance to most of the approved highly potent neutralizing monoclonal antibodies, as well as to the polyclonal antisera induced by infection with the ancestral SARS-CoV-2 and by all the vaccines developed to date (Dejnirattisai, Zhou, Supasa, et al., 2021; Z. Wang, Muecksch, et al., 2021; Wibmer et al., 2021; Zhou et al., 2021). It is generally believed that it was the emergence and rapid spread of VOCs that are largely responsible for the documented cases of SARS-CoV-2 re-infection (Kustin et al., 2021; Naveca et al., 2021; Ortloff, Harsch 2021; Tegally et al., 2021). Specifically, post-vaccination antisera from Moderna and BioNTech vaccinees were 6.5–40-fold less potent against the B.1.351 VOC, but most typically neutralization was reduced 3–8-fold compared to that of the Wuhan-1 SARS-CoV-2 strain (Abdool Karim, de Oliveira 2021; Garcia-Beltran et al., 2021). So far, qualitative and quantitative data on the VOC neutralization by Sputnik V-induced antisera have been very limited (Gushchin et al., 2021; Ikegame et al., 2021).

Notably, in contrast to Moderna mRNA-1273, BioNTech BNT162b2, and Janssen Ad26.COV2.S vaccines that were designed to present the SARS-CoV-2 Spike protein in its pre-fusion conformation, Sputnik V is based on a native Spike protein lacking such modifications. This, in turn, may underlie distinct immune responses, upon cross-platform comparisons, and warrants in-depth analysis.

In this study, we aimed to determine i) whether Sputnik V vaccination is efficient in inducing durable memory B cell (MBC) immunity, ii) whether MBCs are capable of secreting virus-neutralizing antibodies, and if so, iii) whether Sputnik-induced serum antibodies are protective against one of the most neutralization-resistant SARS-CoV-2 viral variants B.1.351 or not. To date, these issues have not been explored in detail and addressing these research gaps should be instrumental for the development of next-generation SARS-CoV-2 vaccines.

## Results

Twenty-two healthy subjects were recruited during the winter/spring of 2021 to receive two doses of the Gam-COVID-Vac (Sputnik V) vaccine (Figure 1a, Supplementary figure 1). The demographic characteristics of this cohort are provided in Supplementary Table 1. The age of the volunteers ranged from 25 to 70 years (median age, 60.0 years; an interquartile range (IQR), 49.8–63.0 years; 64% female).

**Figure 1.**
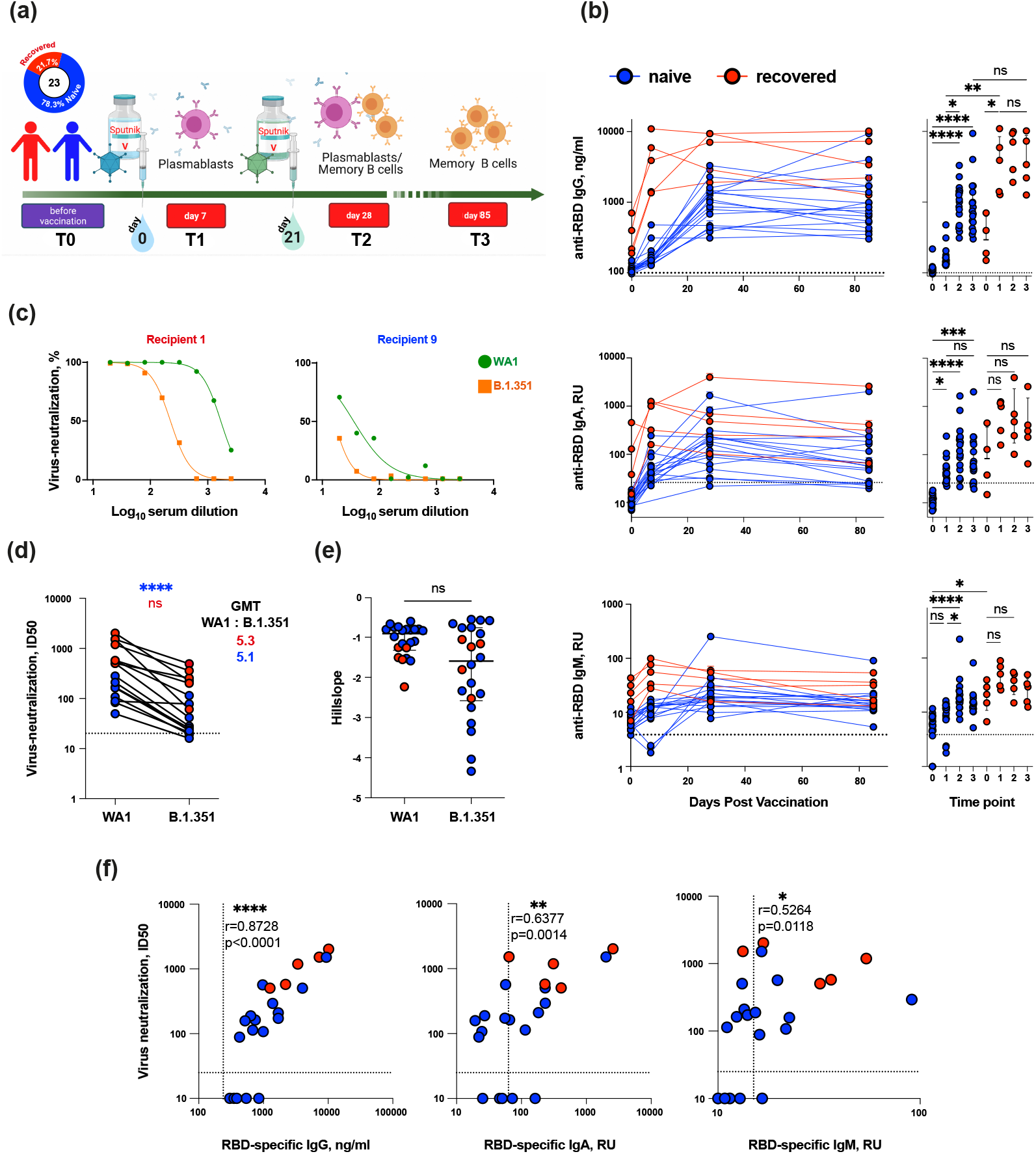
Virus-binding and virus-neutralizing activity of sera from Sputnik V-vaccinated individuals. (a) Study design. (b) Serum anti-RBD IgG (top row), IgA (middle row) or IgM (bottom row) levels for all Sputnik V-vaccinated individuals, measured by ELISA. IgA and IgM levels are shown as a relative units (RU) against a standard convalescent serum. (c) Representative neutralization curves of vaccine-induced sera from one recovered and one naïve individual at T3 time point. (d) Paired analysis of neutralization titers (ID50) against Wuhan strain (WA1) and B.1.351 at T3 time point. (e) Analysis of hillslopes of virus neutralization curves for sera from vaccinated individuals. (f) Spearman’s correlation between serum virus neutralization half-maximal inhibitory serum dilution (ID50) values and the serum levels of anti-RBD IgG (left panel), IgA (middle panel) and IgM (right panel).

Five individuals recruited in October-November 2020 had experienced mild COVID-19 symptoms prior to vaccination (53–120 days). Although no virus-containing samples were available for these patients, it must be noted that the SARS-CoV-2 viral lineage B.1.1 was predominant in Moscow at that time and it was only in March 2021 that the first B.1.351-associated infections were reported in Moscow (Gushchin et al., 2021). Before vaccination, no nucleocapsid (N)- or receptor-binding domain (RBD)-specific IgGs were detected in the sera of naïve individuals without prior COVID-19 symptoms. In contrast, all recovered recipients with self-reported COVID-19 symptoms had both N- and RBD-specific IgG prior to vaccination (Supplementary figure 2).

### Serum antibody responses to Sputnik V vaccination

Plasma samples from the vaccinated individuals were tested for IgG, IgA and IgM RBD-specific antibodies using ELISA. Longitudinal analysis of circulating serum antibodies showed that the levels of anti-RBD IgG and IgA increased markedly after vaccination (Figure 1b). In naive vaccine recipients, the RBD-specific IgG levels mainly increased after the second vaccine dose (T1 vs. T2, *P*=0.0183). In recovered vaccine recipients, the levels of RBD-specific IgG were higher at baseline and displayed a more pronounced increase after the first vaccine dose (T0 vs. T1, *P*=0.0305). Both in recovered and naïve vaccine recipients, the serum RBD-specific IgG levels achieved after the second dose remained stable until day 85. A similar trend was observed for RBD-specific IgA antibodies; however, the overall increase was not as strong. The anti-RBD IgM response in vaccinated individuals was low. The increase in RBD-specific IgM levels was most pronounced in naïve individuals at T2 (T0 vs. T2, *P*<0.0001), consistent with the primary nature of their immune response. Taken together, these results are consistent with those of previous reports exploring antibody responses to mRNA vaccines (Bos et al., 2020; Goel et al., 2021).

Next, we investigated if plasma from Sputnik V-vaccinated subjects, with or without prior COVID-19 history, was active in terms of virus neutralization against the wild-type Wuhan strain WA1 and B.1.351 VOC. To address this question, a SARS-CoV-2 Spike-pseudotyped virus-neutralization test (pVNT) was used to analyze the sera collected at T3 (Figure 1c). B.1.351 VOC is known to be one of the most neutralization-resistant viral variants (P. Wang et al., 2021) and encompasses multiple Spike substitutions (of which three, K417N, E484K, and N501Y, are in the RBD). All the 22 serum samples were active against WA1, although neutralization potency varied broadly with ID50 ranging from 49–2021 (geometric mean titer [GMT] 290, Figure 1c-d).

Compared to the sera from the naïve group of vaccinees, samples obtained from the recovered group demonstrated significantly higher neutralization activity against both WA1 (ID50 GMT 913 vs. 149, *P*=0.0019), and B.1.351 (ID50 GMT 190.7 vs. 49.8, *P*=0.0111). Importantly, the GMT values for B.1.351 were 5.1-fold lower than those for WA1 in naïve recipients (*P*=0.0250) (Figure 1d). All T3 sera from the five recovered subjects displayed 90% neutralization of B.1.351 at a 1:20 dilution (Supplementary Figure 3). In contrast, in the naïve group, only one out of the total 17 samples (subject 20) displayed similar neutralization potency. Nonetheless, the undiluted sera could achieve 90% neutralization in all but one of the naïve samples. The neutralization activity in that exceptional sample (subject 22) could not be reliably measured. However, the neutralization titers we measured in the cohort of naïve Sputnik V vaccinees were significantly higher than those reported by Ikegame et al. (Ikegame et al., 2021). Specifically, they were three-fold higher for the ancestral WA1 Spike (ID50 GMT 150; 95% CI - 90.1– 248.5) and seven-fold higher for B.1.351 (ID50 GMT 49; 95% CI - 28.67–83.49). Neutralization curve shape comparison did not reveal significant differences between the Hill slopes for WA1 and B.1.351 (*P*=0.0803) (Figure 1e), indicating that the neutralization abilities of sera towards WA1 and B.1.351 vary more in quantitative rather than in qualitative terms.

Finally, the levels of WA1 virus neutralization were strongly correlated with the level of RBD-specific IgG and modestly correlated with that of IgA and IgM (Spearman’s r=0.8728, *P*<0.0001; r=0.6377, *P*<0.0014; r=0.5264, *P*<0.0118, for IgG, IgA, and IgM, respectively) (Figure 1f), suggesting that IgG antibodies are the most potent neutralizing component of the sera.

Blue and red symbols indicate naïve (n = 17) and recovered (n = 5) participants. Symbols connected by solid lines represent time points considered for each individual. Data are presented as median ⍰ IQR. The dotted lines indicate the threshold for positivity. Asterisks indicate significant difference between groups determined using the Kruskal–Wallis test, *P < 0.05, **P < 0.01, ***P < 0.001, ****P < 0.0001, ns = not significant. GMT, geometric mean titer; ID50, half-maximal inhibitory dilution; IQR, interquartile range; RBD, receptor-binding domain; RU, relative units.

### Total and RBD-specific plasmablast response

One of the earliest manifestations of the B cell response is the emergence of circulating total and antigen-specific plasmablasts, which peak around the seventh day post-immunization (Wrammert et al., 2008). Plasmablasts were defined here as CD3^-^CD16^-^CD19^+^IgD^-^CD27^hi^CD38^hi^ cells (Wrammert et al., 2008) (Figure 2a, left panel).

**Figure 2.**
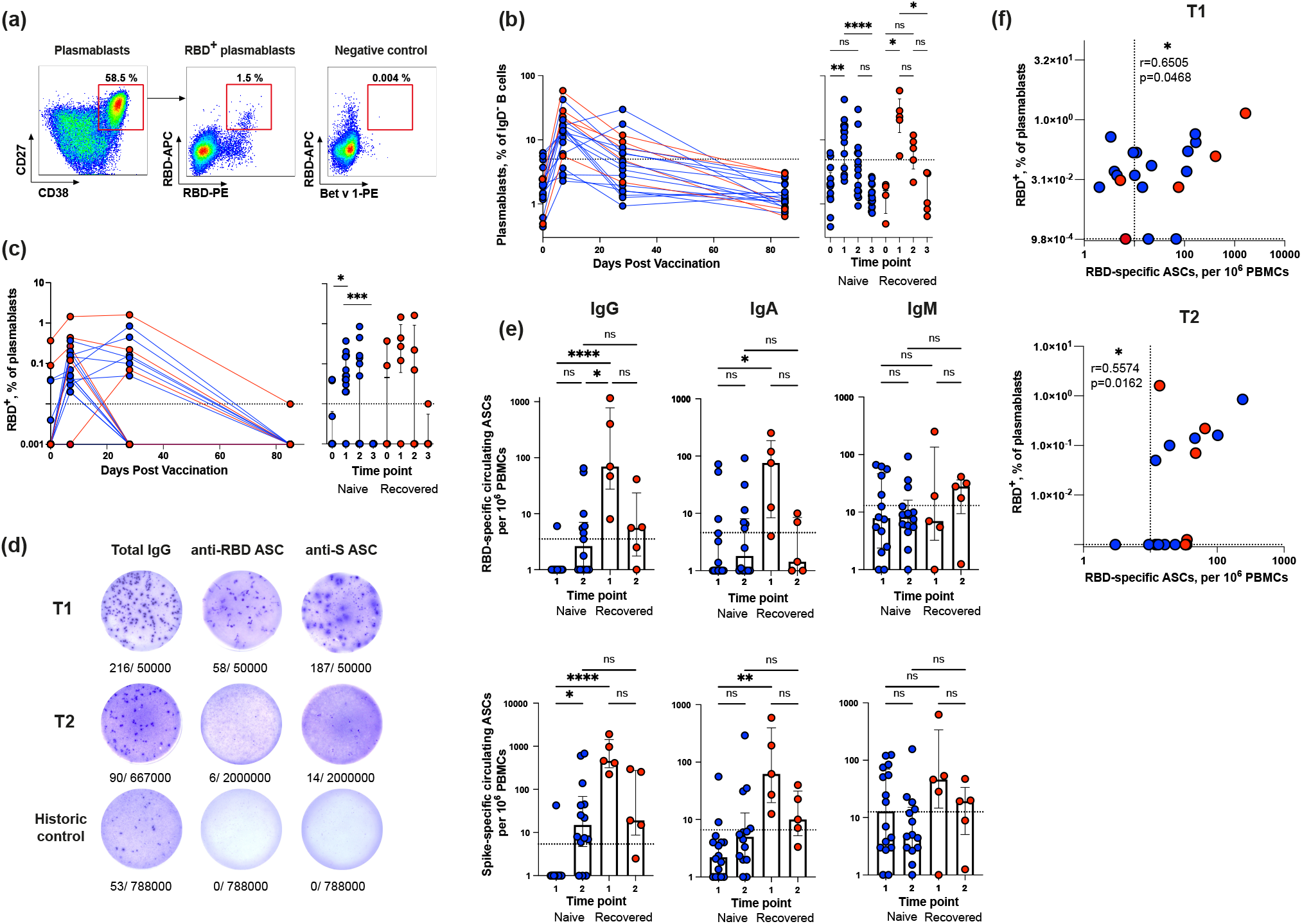
Quantification of total and RBD-specific plasmablasts in blood samples from Sputnik V-vaccinated individuals. (a) Representative flow cytometry dot plots showing the gating strategy for measuring the percentage of total (left panel) and RBD+ (middle panel) plasmablasts. As a negative control, sample stained with an irrelevant protein Bet v 1 is shown (right panel). Numbers inside the plots indicate the percentage of events specific to respective gates. (b-c) Dynamic changes in total (b) and RBD+ (c) plasmablast frequencies in samples collected at different time points. (d) Representative ELISpot images for measuring the frequencies of circulating total (left), anti-RBD (middle) or anti-S (right) IgG ASCs. The wells shown contained 106 PBMCs obtained from vaccinated individuals. Numbers below the wells represent the frequencies of ASCs relative to the total number of cells in the well. (e) Frequencies of circulating IgG (left column), IgM (middle column) or IgA (right column) ASCs specific for RBD (upper row) or S (bottom row) antigens per 106 PBMCs collected from vaccinated individuals at different time points. The dotted lines indicate the threshold for positive antigen-specific ASC responses. (f) Spearman’s correlation between RBD-specific (IgG + IgA + IgM) ASCs and the levels of RBD+ plasmablasts at T1 (left) and T2 (right) time points.

Before vaccination, the plasmablast frequencies were the same as those in normal donors but increased markedly after the primary immunization (T0 vs. T1 *P=*0.0076 and *P*=0.0289 for naïve and recovered individuals, respectively) (Figure 2b). Booster immunization resulted in an insignificant increase in the percentage of total plasmablasts compared to the baseline level. Positive plasmablast response was detected in 81% (17/21) of cases at T1 and only in 40% (8/20) of cases at T2. Subsequently, as can be seen from the measurements at T3, plasmablasts completely disappeared from the circulation.

The dynamics of SARS-CoV-2-specific plasmablasts during vaccination were of special interest to us. Since virus-neutralizing antibodies are known to predominantly target the RBD (Dejnirattisai, Zhou, Ginn, et al., 2021; Yang, Du 2021), when detecting antigen-specific B cells, we focused on detecting RBD-binding (RBD^+^) cells. Antigen-specific plasmablasts were detected by double staining using RBD-PE and RBD-APC (Figure 2a, middle panel). As a negative control, we used the samples stained with an irrelevant PE-labeled protein Bet v 1, which is the major birch allergen (Figure 2a, right panel) (none of the study participants had birch pollen allergy). Based on this negative control, the cut-off value for RBD-binding plasmablasts was set at 0.01%. In contrast to COVID-19 patients sampled in the acute phase of the disease (Byazrova et al., 2021), the frequencies of RBD^+^ plasmablasts in vaccinated individuals were low (Figure 2c). The RBD^+^ plasmablast response was detectable in 18 subjects (86%) after the first dose of Sputnik V and only in eight participants (40%) after the booster immunization. In only one subject from the recovered subgroup, it exceeded 1%, which is still significantly lower than what was observed in patients with moderate COVID-19 (Byazrova et al., 2021).

Since plasmablasts are antibody-secreting cells (ASCs), it is logical to detect and enumerate them using an enzyme-linked immunosorbent spot (ELISpot) assay. Representative ELISpot images of circulating Spike- and RBD-specific IgG ASCs are presented in Figure 2d. Historic control wells showed only rare spontaneous total ASCs and no SARS-CoV-2-specific ASCs. The magnitude of the IgG ASC response was the largest in the recovered group after the first vaccine dose, when RBD- and S-specific IgG ASCs were detected in all vaccine recipients (median 69, IQR 27.33–772.8 and median 459, IQR 318– 1439 for RBD- and S-specific ASCs, respectively) (Figure 2e). In contrast, no RBD- or S-specific IgG ASCs were found in all but one naïve subject after the first dose. After the second vaccination, 7 and 11 naive participants (n=15) had RBD- and S-specific IgG ASCs above the baseline, respectively. The fact that after the first vaccine dose, the frequencies of anti-RBD IgG ASCs were higher in SARS-CoV-2-recovered individuals than in individuals in the naïve group is consistent with the idea that, in the former cohort, this increase is due to the re-activation of MBCs. The SARS-CoV-2-specific IgM responses mediated by the circulating ASCs were generally lower in their magnitude than the IgG responses and we did not observe significant differences between the naive and recovered samples, nor between the T1 and T2 time points.

Plasmablasts are a heterogeneous population of cells that are usually subdivided into early and later plasmablasts based on their ability to express the B cell receptor (BCR) on their surface and secrete antibodies (Sanz et al., 2019). For the most part, plasmablasts express membrane bound BCR and simultaneously secrete antibodies. However, surface BCR expression is more a characteristic of early plasmablasts, while antibody-secreting capacity is more associated with later plasmablasts. Accordingly, we found a modest correlation between the frequencies of RBD^+^ plasmablasts and RBD-specific Ig (IgG + IgA + IgM) circulating ASCs (Spearman’s r=0.6505, *P*=0.0468 at T1; r=0.5574, *P*=0.0162 at T2) (Figure 2f).

Results are shown for individual samples (symbols) from naïve (n = 17) and recovered (n = 5) recipients. Data are presented as median ⍰ IQR. Asterisks indicate significant difference between groups determined using the Kruskal–Wallis test, *P < 0.05, **P < 0.01, ***P < 0.001, ****P < 0.0001, ns = not significant. ACS, antibody-secreting cell; IQR, interquartile range; RBD, receptor-binding domain.

### SARS-CoV-2-specific memory B cell response after Sputnik V vaccination

To investigate whether Sputnik V-vaccinated individuals developed antigen-specific MBCs, we used two complimentary approaches, namely, flow cytometry of RBD-binding circulating (Dan et al., 2021; Hartley et al., 2020) and quantification of SARS-CoV-2-specific ASCs induced by *in vitro* interleukin 21 (IL-21)/CD40L stimulation (Byazrova et al., 2021; Yao et al., 2021). The RBD^+^ MBCs were defined as CD19^+^CD27^+^CD38^-^ and double-positive for the fluorescently labeled RBD-PE and RBD-APC following exclusion of IgD^+^ B cells (Figure 3a). The frequencies of RBD^+^ MBCs measured at different time points are shown in Figure 3b. Even before vaccination, the recovered individuals had a noticeable number of RBD^+^ MBCs, which was higher than that of naive subjects (*P*=0.0362), and above the level of the negative control defined by staining with an irrelevant Bet v 1 protein (0.01%). Until day 85, the level of RBD^+^ MBCs in recovered individuals remained on average stable. In naïve individuals, baseline frequencies of RBD+ MBCs were observed at T0 and T1. At T2, RBD^+^ MBCs were detected above the threshold in 37.5% of naïve individuals, displayed a further increase at T3 (T1 vs. T3, *P*=0.0023), and approached the level observed in recovered individuals.

**Figure 3.**
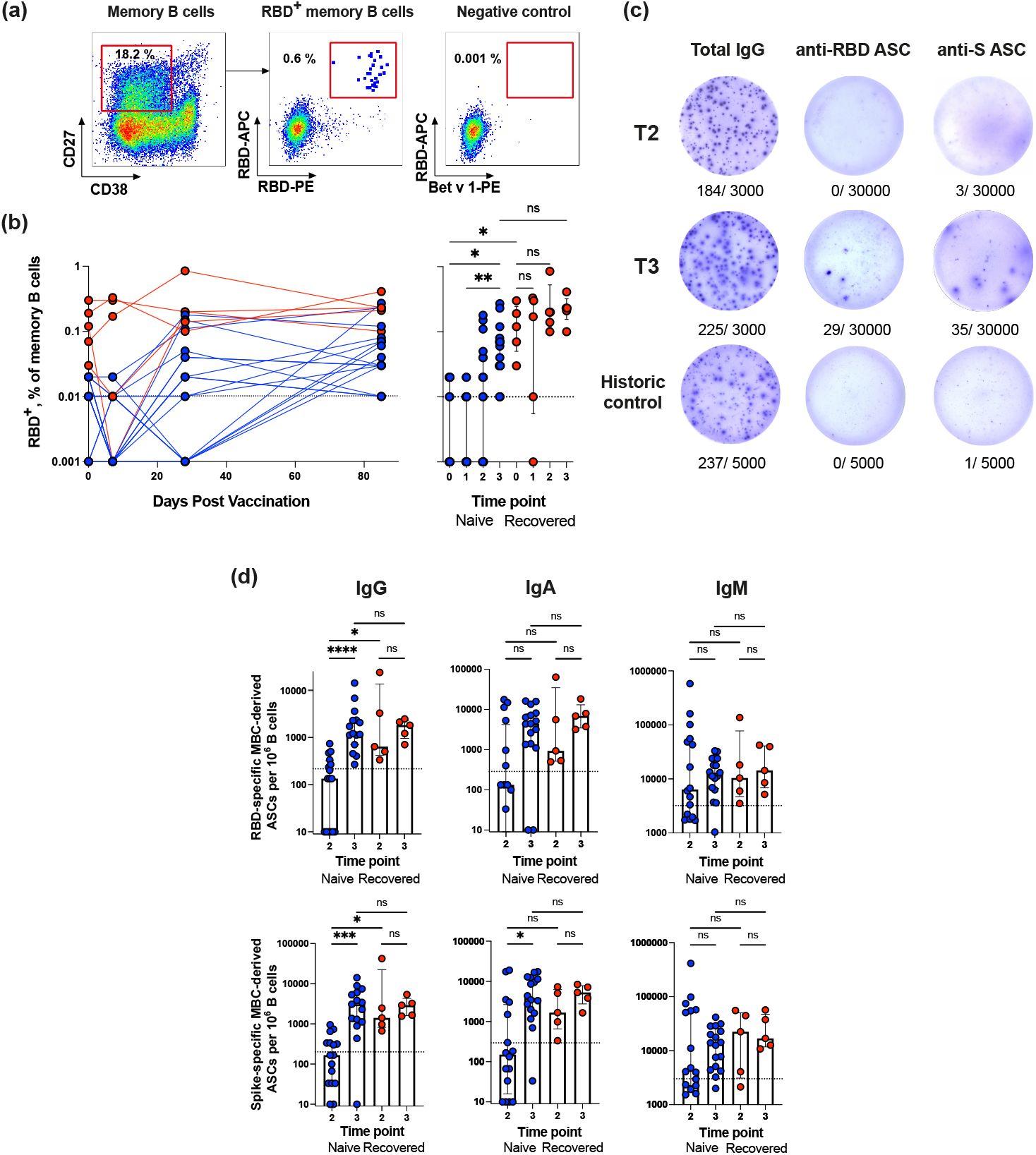
Analysis of the MBC response in Sputnik V-vaccinated individuals. (a) Representative flow cytometry dot plots showing double discrimination of RBD+ MBCs. Numbers inside the plots indicate the percentage of events specific to the respective gates. (b) RBD+ MBCs as a percentage of all memory B cells (CD19+CD27+CD38-IgD-). (c) Representative ELISpot showing SARS-CoV-2-specific MBC-derived ASCs. Purified B cells were stimulated with IL-21/CD40L for 7 days and then incubated in ELISpot plates for 16 h to detect ASCs secreting total (left column), RBD-(middle column) or S-specific (right column) IgG at T2 time point (upper row), T3 (middle row) or in historic control samples (bottom row). The numbers indicated below the wells represent positive dots and the total number of cells in the well. (d) RBD-(upper row) and S-specific (bottom row) MBC-derived ASCs per 106 B cells from blood samples of naïve (n = 17) or recovered (n = 5) vaccinated individuals at different time points. Data for IgG (left column), IgM (middle column) and IgA (right column) ASCs are presented. The dotted lines indicate the threshold for a positive antigen-specific ASC response calculated with pre-pandemic samples per 106 B cells.

Measurements of the SARS-CoV-2-specific circulating MBC numbers were supplemented with a more functional ELISpot assay. In contrast to plasmablasts, MBCs are resting cells and do not secrete antibodies without stimulation. For MBC activation and induction of antibody secretion, immunomagnetically purified B cells were stimulated *in vitro* with IL-21/CD40L. These conditions mimic the germinal center environment in which B cells differentiate into plasma cells (Ding et al., 2013). After stimulation for 7 days, the frequencies of S- and RBD-specific ASCs were evaluated using an ELISpot assay. As shown in the representative ELISpot images (Figure 3c), our protocol for polyclonal B cell activation was highly efficient and resulted in the secretion of both total and SARS-CoV-2-specific antibodies. In pre-pandemic control samples, the frequencies of SARS-CoV-2-specific IgG ASCs were below 200 and 215 spots per million B cells were seeded in Spike- and RBD-coated wells. These values served as a cut-off for positivity. The MBC-derived ASC numbers were measured at two time points, 28 (T2) and 85 (T3) days after the first dose of the vaccine, when MBCs become detectable (T2) and undergo maturation (T3).

MBC-derived IgG ASCs displayed the strongest response. At T2, in all recovered (5/5) and in some naive subjects (7/17) the SARS-CoV-2-specific MBC-derived ASC numbers were above the baseline; however, recovered subjects had a higher level of ASCs than naïve subjects (*P*=0.034 and *P*=0.0373 for RBD- and S-specific ASCs, respectively) (Figure 3d). At T3, the numbers of ASCs in recovered vaccinees remained stable, while in naïve subjects they increased 14-fold and approached the recovered group level (in the naïve group T2 vs. T3, *P*<0.0001 and *P*=0.0002 for RBD- and S-specific ASCs, respectively). On average, at the peak of the response, approximately 3,000 RBD-specific IgG ASCs per million B cells (0.3% of total B cells) were detected, which was similar to the number of RBD^+^ MBCs detected using flow cytometry in the recovered group (median 0.23%) (Figure 3b). The dynamics of the RBD-specific IgG ASC response was also consistent with the flow cytometry data. RBD-specific ASCs constituted 8–100% of all S-specific ASCs (Figure 3d), suggesting that most of the MBC-derived antibodies were directed against the RBD.

The frequencies of IgA MBC-derived ASCs targeting the Spike and RBD followed a similar pattern, namely, recovered subjects responded more quickly than naïve subjects, but the responses of both groups were roughly comparable at T3. Although the anti-SARS-CoV-2 IgM ASC responses were above the threshold, no significant differences were found between the naive and recovered groups, or between T2 and T3. Thus, our results indicate that in naive recipients, the maximum number of vaccine-induced MBCs is reached only 85 days after the first vaccination, and in terms of kinetics, this process significantly lags behind the formation of serum antigen-specific antibodies. Thus, on day 85 after the first dose of Sputnik V, almost all the vaccinees developed RBD- and S-specific MBCs.

Results are shown for individual samples (symbols) from naïve (n = 17) and recovered (n = 5) recipients. Data are presented as median ⍰ IQR. Asterisks indicate significant difference between groups determined using the Kruskal–Wallis test, *P < 0.05, **P < 0.01, ***P < 0.001, ****P < 0.0001, ns = not significant. ASC, antibody-secreting cell; IQR, interquartile range; RBD, receptor-binding domain.

### MBC-derived SARS-CoV-2 antibody reactivity

We then explored the overall level of virus-binding antibodies that MBCs from the vaccinated subjects could provide, and their virus-neutralizing activity. To address these two questions, we used 7-day cultures of IL-21/CD40L-stimulated B cells as a source of MBC-derived antibodies and performed ELISA using the culture supernatants on RBD-precoated plates. The stimulated MBCs from all recovered individuals secreted a significant amount of anti-RBD IgG both at T2 and T3 (Figure 4a). In the naïve group at T3, the level of anti-RBD IgG secretion was five- to six-fold reduced compared to that of the recovered group. Stimulated MBCs produced anti-RBD IgA and IgM less efficiently than IgG, and the difference between samples from naïve and recovered subjects was less pronounced.

**Figure 4.**
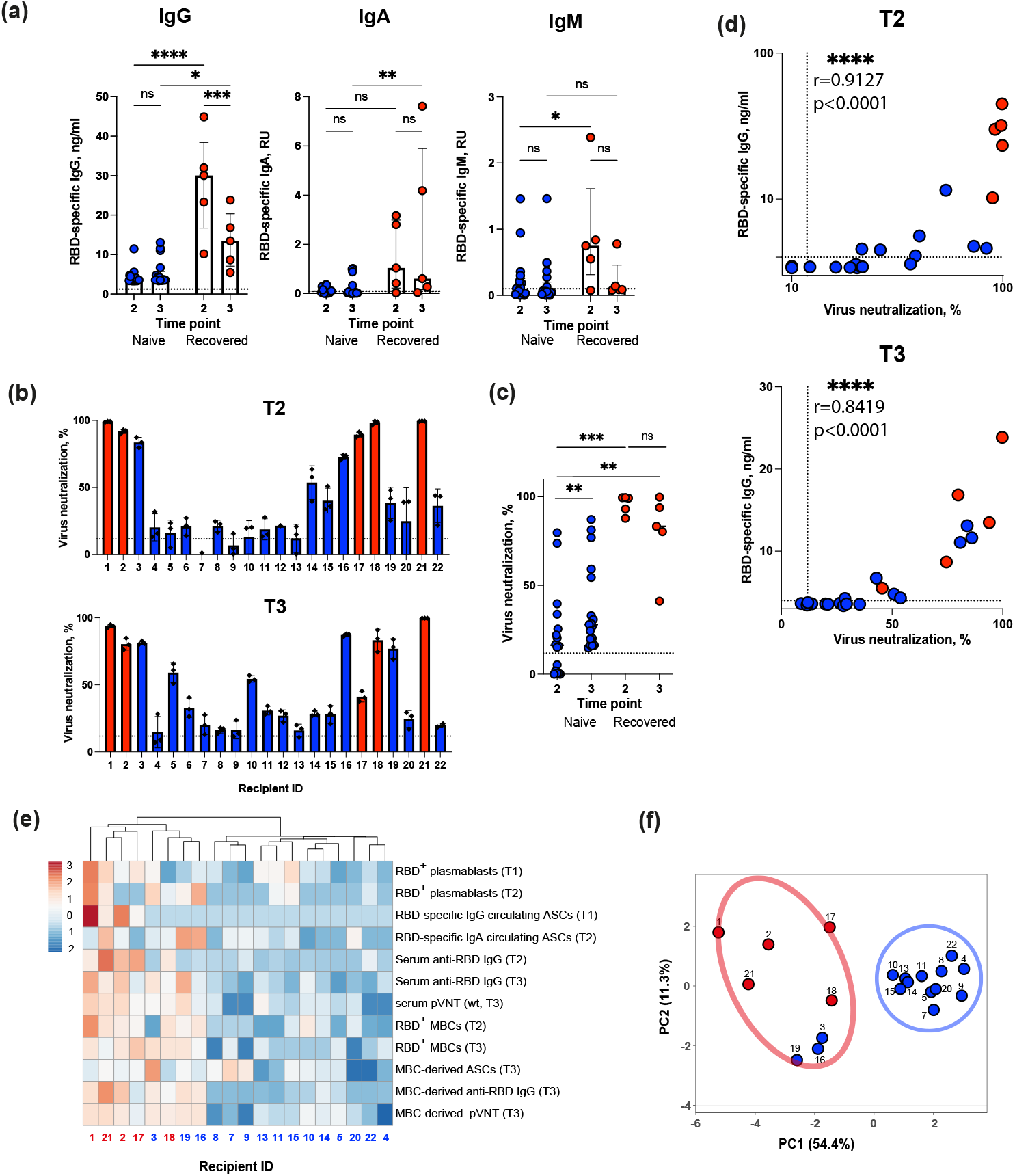
Analysis of MBC-derived antibody response in supernatants of CD40L/IL-21 stimulated B cells from Sputnik V-vaccinated individuals. (a) Production of RBD-specific IgG (left panel), IgA (middle panel) or IgM (right panel) in cultures of IL-21/CD40L-stimulated B cells from Sputnik V-vaccinated individuals evaluated using ELISA. (b) Virus-neutralizing activity of antibodies derived from the cultures of CD40L/IL-21-stimulated B cells for individual vaccine recipients at T2 (upper panel) or T3 (bottom panel) time point. The results of three separate experiments each with two technical repeats are presented for all vaccine recipients. (c) Summary of virus-neutralizing activities of antibodies derived from cultures of CD40L/IL-21-stimulated B cells from vaccinated individuals. (d) Spearman’s correlation between virus neutralization (%) and the levels of anti-RBD IgG in supernatants of IL-21/CD40L-stimulated B cells obtained from Sputnik V-vaccinated individuals at T2 (upper panel) and T3 (bottom panel) time points. (e) Heatmap and hierarchical clustering of Sputnik V recipients. Columns denote Sputnik V recipients. Rows correspond to immune response variables. Dendrograms on the top illustrate the clustering of Sputnik V recipients. Immune response measurement values are color-coded according to the key shown in the upper left. (f) Principal component analysis of Sputnik V recipients. Recipient IDs are shown. Two distinct clusters are indicated by the ovals.

Next, we tested the virus-neutralizing activity of MBC-derived antibodies using a pVNT assay. Since the concentration of antibodies in the supernatants was approximately two orders of magnitude lower than that in the plasma, undiluted supernatants were used. MBC-derived antibodies from recovered individuals inhibited pseudovirus entry in the range of 37–99% at both time points T2 and T3 (Figure 4b). At T2, 12/17 individuals from the naïve group were also responders in the pVNT assay (median neutralization 23.3%, IQR 16.84–68.1%). At T3, all naïve subjects demonstrated virus neutralization above the threshold, and the neutralization activity of MBC-derived antibodies also increased (T2 vs. T3, *P*=0.004) (Figure 4c). For stimulated MBCs, strong correlations were observed between the virus-neutralization of MBC-derived antibodies and the level of *in vitro* anti-RBD IgG secretion both at T2 (Spearman’s r=0.9127, *P*<0.0001) and T3 (Spearman’s r=0.8419, *P*<0.0001) (Figure 4d). Based on these findings, it may be possible to assess the quality of antibodies that MBCs will produce upon re-exposure to an antigen.

We observed that naïve and recovered vaccine recipients differed in several key parameters of humoral and B cell immunity. To investigate if study participants could be divided into subgroups, we carried out hierarchical cluster and principal component analyses. Based on the 12 humoral and B cell response measurements, these analyses, which included 15 naïve and five recovered vaccine recipients indicated the existence of two well-separated clusters (Figure 4e, f). The first compact cluster encompassed exclusively naïve vaccine recipients; the second, somewhat loose cluster included all the recovered and three naïve vaccine recipients. Thus, when several humoral and B cell parameters were taken into consideration, the distinct immune profiles of the naïve and recovered vaccine recipients became clearly apparent. Most interestingly, our comprehensive B cell profiling analysis has uncovered the existence of two categories of vaccine recipients, namely, the high- and low-responders. This highlights the underappreciated heterogeneity of the human immune response to Sputnik V, thereby warranting a systematic identification of predictors and modifiers of this response.

Results are shown for individual samples (symbols) from naïve (n = 17) and recovered (n = 5) recipients. Data are presented as median ⍰ IQR. Asterisks indicate significant difference between groups determined using the Kruskal–Wallis test, *P < 0.05, **P < 0.01, ***P < 0.001, ****P < 0.0001, ns = not significant. ASC, antibody-secreting cell; IQR, interquartile range; RBD, receptor-binding domain.

## Discussion

Presently, Sputnik V is used in several countries including Russia, Argentina, India, and Brazil. Nonetheless, the protective properties of this vaccine have been debated and the information available has been somewhat ambivalent (Bucci et al., 2021; Gushchin et al., 2021; Ikegame et al., 2021; Logunov et al., 2021; 2020; Rossi et al., 2021). In the present study, a comprehensive analysis of B cell immunity was performed in a cohort of 22 vaccinated subjects, among whom 5 had previously recovered from mild COVID-19. Several parameters were analyzed, namely, (i) the serum antibody titers to RBD, (ii) activity of serum antibodies in the pVNT assay with wild-type SARS-CoV-2 and its mutant variant B1.351, (iii) detection of RBD-specific plasmablasts and MBCs, (iv) enumeration of circulating and MBC-derived ASCs, and the (v) virus-binding and -neutralizing activity of MBC-derived antibodies. Most of these parameters were measured during vaccination with the final time point being 2 months after the first vaccine dose. This analysis has allowed us to compare the dynamics and magnitude of B cell immune responses in naïve and COVID-19-recovered vaccine recipients, which reflects the real-world epidemiological situation.

Taken together, our results demonstrate faster and more robust B cell responses to Sputnik V in COVID-19-recovered individuals than in individuals without prior infection. Considering many parameters, recovered individuals achieved the maximum immune response after the first vaccine dose. This is in line with the results obtained in mRNA vaccine studies (Ebinger et al., 2021; Goel et al., 2021; Krammer et al., 2021; Stamatatos et al., 2021; Z. Wang, Muecksch, et al., 2021). Importantly, we, for the first time, demonstrate this effect at the level of both plasmablasts and MBCs rather than just via quantification of antibody responses and assessment of their neutralization activity. Combined with previously reported data (Rossi et al., 2021; Sasikala et al., 2021), our results indicate that a single dose of the adenovirus-based anti-SARS-CoV-2 vaccine, like mRNA-based vaccines, may be sufficient for protective immunity, when used in subjects with pre-existing immunity to SARS-CoV-2.

Among naive vaccine recipients, the response to Sputnik V vaccination was overall slower than that of recovered vaccine recipients, and a fraction of vaccinees who received both doses of Sputnik V never displayed the concentration and neutralization potency of antibodies that would match the levels observed in recovered individuals. Importantly, after the second dose of Sputnik V, the antisera of naïve vaccine recipients showed virus-neutralizing activity against the wild-type virus in all but one individual (recipient 22). On average, the antisera from naïve Sputnik V recipients were less potent in terms of their neutralizing ability as compared to the antisera from the mRNA vaccines (Anderson et al., 2020; Liu et al., 2021; Wu et al., 2021), yet they were on par with the numbers reported for other adenovirus-based vaccines (Folegatti et al., 2020; Zhu et al., 2020).

Furthermore, we wanted to investigate if Sputnik V vaccination may efficiently neutralize mutant SARS-CoV-2 variants. Whereas detailed studies are available for the Pfizer and Moderna vaccines, data for Sputnik V are limited. Recently, it was reported that antisera from naïve Sputnik V recipients were 3.1-, 2.8-, and 2.5-fold less potent against the B.1.351, B.1.1.248, and B.1.617.2 variants, respectively (Gushchin et al., 2021). Another recent study references a median 6.1-fold reduction in the GMT against B.1.351 (Ikegame et al., 2021) with a notable comment that when extrapolated to full serum strength, half of the serum samples failed to achieve an ID80 and only one out of 12 achieved an ID90 against B.1.351. In this study, we found that the GMT of sera from recovered and naïve recipients exhibited five-fold reduction against B.1.351 compared to that against the wild-type virus in a pVNT assay, but the neutralizing activity of all but one samples was sufficient to achieve an ID90 against B.1.351 when extrapolated to full serum strength.

The neutralizing potency of the antisera from recovered subjects against the wild-type variant was more than four-fold higher than that of the naive group. Predictably, these samples were also much more active against the B.1.351 variant. This is in excellent agreement with the data reported for mRNA vaccines (Goel et al., 2021) and is indicative of the higher level of protection against emerging viral variants in the SARS-CoV-2 pre-exposed vaccinees.

It is interesting to compare the B cell immunity elicited by Sputnik V vaccination with that induced by natural SARS-CoV-2 infection. The dynamics of plasmablast numbers is of special interest because it can be used as a predictor of successful humoral immunity (Appanna et al., 2016). We found low levels of RBD^+^ plasmablasts and circulating RBD-specific ASCs in vaccinated naive individuals compared to those observed in the acute phase in COVID-19 patients (Byazrova et al., 2021; Kuri-Cervantes et al., 2020). Perhaps these differences are associated with the extrafollicular pathway of B cell activation in the acute phase of COVID-19, which is characterized by massive plasmablast expansion (Woodruff et al., 2020). It has been suggested that this pathway may contribute to the pathogenesis of acute COVID-19. From this standpoint, the modest plasmablast response observed during vaccination can be viewed as beneficial.

Unlike the plasmablast response, the MBC response was well-pronounced, and the MBC numbers observed are comparable to those found in acute COVID-19 patients. This is important because vaccine-induced MBCs are known to be central to the longevity of immune memory and are among the first cells to produce massive amounts of antibodies upon antigen re-encounter. Although the MBC numbers are informative descriptors of B cell immunity, the functional activity of the antibodies that will be produced during secondary immune responses is key to our understanding of vaccine-induced protection. To address this question, antigen-specific MBCs were isolated followed by single-cell sequencing of Ig genes and expression of recombinant MBC-derived antibodies (Abayasingam et al., 2021; Dejnirattisai, Zhou, Supasa, et al., 2021; Scheid et al., 2021; Z. Wang, Schmidt, et al., 2021). Alternatively, antibody secretion can be induced in cultures of polyclonally stimulated B cells. Seven-day cultures of IL-21/CD40L-stimulated MBCs isolated from the blood of Sputnik V vaccinees secreted anti-RBD IgG at approximately the same level as MBCs from acute COVID-19 patients.

One of the most interesting observations we made is that the ancestral variant of SARS-CoV-2 could be neutralized by MBC-derived antibodies from the vast majority of Sputnik V vaccinees. Two months following vaccination, the differences between the naïve and recovered vaccine recipient cohorts in terms of MBC numbers and their ability to differentiate into ASCs that secrete RBD-specific and virus-neutralizing antibodies, became less pronounced, unlike the differences in the neutralizing titers of serum antibodies. This indicated the gradual maturation and continued evolution of the MBC population. Thus, we show that *in vitro* stimulation of virus-specific MBCs can considerably extend the traditional serological analysis of vaccinated donors. This will allow the study of the dynamics and longevity of antigen-specific MBCs in the course of infection and vaccination. Using this approach, we provide experimental evidence indicating that both recovered and naïve vaccinees accumulate similar numbers of virus-specific MBCs at 2 months after the second dose of Sputnik V. Upon antigen stimulation, these cells differentiate into ASCs that secrete virus-specific antibodies, of which a significant proportion is virus-neutralizing. Based on these data, we conclude that the MBCs elicited by Sputnik vaccination, both in their number and productivity, are comparable to those generated during natural infection.

As in the scenario of natural infection, some individuals, classified as low responders, fail to mount robust immune response upon Sputnik V vaccination. The reasons underlying the poor vaccine immunogenicity in these subjects are presently unknown but are likely related to individual features of the immune system. However, our study group was rather limited; 2 months after vaccination, one recipient with the lowest levels of virus-neutralizing antibodies and other indicators of poor B cell immunity developed PCR-confirmed COVID-19 (to be published). Currently, the minimum levels of serum virus-binding and virus-neutralizing activity that are protective against vaccine breakthrough infections remain to be determined. Hence, additional studies are required to establish this minimum level of immunity.

## Materials and Methods

### Volunteers

A cohort of 22 Sputnik V recipients was enrolled in December 2020 at the National Research Center Institute of Immunology of The Federal Medical Biological Agency of Russia. None of the participants were pregnant, immunodeficient, or receiving immunosuppressive treatment. Subjects were immunized by intramuscular injection into the deltoid muscle with a 21-day interval between the doses. All subjects received two doses of Gam-COVID-Vac (Sputnik V) vaccine. None of the volunteers had experienced serious adverse events after vaccination. Written informed consent was obtained from each of the study participants before performing any study procedures. The study protocol was reviewed and approved by the Medical Ethical Committee of Institute of Immunology (#12-1, December 29, 2020).

### Blood sample collection and processing

Whole-blood samples were collected into heparinized vacutainer tubes (Sarstedt, Cat. No. 04.1927) four times: one day before vaccination, on day 7 after the first and the second doses of vaccine, and on day 85 from the start of vaccination (T0, T1, T2, and T3 time points, respectively) (Supplementary Fig. 1).). PBMCs were isolated by density gradient centrifugation. Plasma samples were stored at -80°C. B cells were purified from PBMCs by negative selection using the Dynabeads Untouched human B cells kit (Thermo Fisher Scientific, Cat. No. 11351D).

Immunomagnetically separated B cells were cultured in complete DMEM/F12 medium (Cat. No. C470p) supplemented with 10% FBS (Cat. No. SV30160.03), 2 mM L-glutamine (Cat. No. F032), 24 μg/mL gentamicin (Cat. No. A011p), 1 mM sodium pyruvate (Cat. No. F023), and 10 mM HEPES (Cat. No. F134) (all from Paneko). To obtain MBC-derived antibody-secreting cells (ASCs), B cells were stimulated with 25 ng/mL interleukin-21 (IL-21; PeproTech, Cat. No. 200-21) in the presence of mitomycin-treated feeder A549 cells stably expressing CD40L (A549-CD40L, 1 × 10^5^ cells/well) for 7 days at a density of 5 × 10^3^ B cells/well in 96-well plates at 37°C in 5% CO_2_. Stimulated B cells were harvested 7 days later and used in ELISpot assay. In parallel, supernatants from IL-21/CD40L-stimulated B cells were also collected on day 7 of co-culture for measuring the levels of secreted antibodies in ELISA or virus neutralization assays.

### ELISA

The level of SARS-CoV-2 receptor binding domain (RBD)-specific antibodies was measured using ELISA Quantitation Kit (Xema Co., Cat. No. K153G). Plasma samples from vaccinated individuals or supernatants from IL-21/CD40L stimulated B cells were 2-fold serially diluted from 1:20 to 1:12500 and 1:2 to 1:200 respectively in blocking buffer. Plates were incubated with samples for 1 hour at room temperature. After washing, the plates were additionally incubated for 1 hour with anti-human IgG, IgM or IgA secondary antibody conjugates with horseradish peroxidase (Jackson Immuno Research, Cat. No. 109-036-088, 109-035-129, and 109-035-011) diluted 1:5,000 in blocking buffer. ELISA plates were washed 7 times and developed for 10 min with 100 μL of TMB chromogen solution. The reaction was stopped by adding 50 μl 1 M H_2_SO_4_ and optical density at 450 nm was measured using the iMark microplate absorbance reader (Bio-Rad, Cat. No. 1681130). Each sample was measured in triplicate. To determine the concentration of IgG, a serial dilution of anti-SARS-CoV-2 RBD-specific human monoclonal antibody iB12 was included on each plate, a calibration curve was built and IgG levels were calculated (μg/mL). When determining the levels of IgM and IgA, we used high-titer convalescent serum as a standard and antibody levels were expressed as relative units (RU).

### Flow cytometry

Freshly isolated PBMCs were stained with the following antibodies: CD3 FITC (clone TB3), CD16 FITC (clone LNK16), CD19 PE (clone LT19), CD27 PECy5.5 (clone LT27), CD38 PECy7 (clone LT38) (all were produced in-house earlier (Khvastunova et al., 2015); CD14 FITC (clone MEM-15, Exbio, Cat. No. ED7028); anti-human IgG APC (clone M1310G05, Biolegend Cat. No. 410720) and anti-human IgM APC-Fire750 (clone MHM-88, Biolegend Cat. No. 314546). RBD-specific B cells were detected using double staining with phycoerythrin- and allophycocyanin-labelled RBD (RBD-PE and RBD-APC). Production of recombinant RBD (isolate Wuhan-Hu-1) or Bet v 1 conjugated to PE or APC was described earlier (Byazrova et al., 2021). Cells were analyzed on a CytoFLEX S flow cytometer (Beckman Coulter). Up to 10 × 10^6^ cells were acquired per sample. Data were analyzed using FlowJo Software (version 10.6.1., Tree Star).

### ELISpot assay

Quantification of SARS-CoV-2-specific ASCs was performed by enzyme-linked immunosorbent spot (ELISpot) assay as described previously (Byazrova et al., 2021). Briefly, sterile clear 96-well Multiscreen HTS Filter Plates with 0.45 μm pore size hydrophobic polyvinylidene difluoride membrane (Merck Millipore, Cat. No. MSIPS4510) were stripped with 70% ethanol for 2 min, washed and coated with 10 μg/mL of recombinant RBD or native ectodomain S protein from SARS-CoV-2 (isolate Wuhan-Hu-1). In-house production of recombinant SARS-CoV-2 proteins was described earlier (Shomuradova et al., 2020). To capture the total immunoglobulin (IgG, IgM or IgA) produced by ASCs, wells were coated with 10 μg/ml of rabbit anti-human IgG or IgM (R&D Systems, Cat. No. SELB002, SELB003), or goat anti-human IgA antibodies (SouthernBiotech, Cat. No. 2050-01).

Freshly purified PBMCs or IL-21/CD40L stimulated B cells were used for quantification of circulating ASCs or MBC-derived ASCs, respectively. Cells were resuspended in complete DMEM/F12 medium and plated at a density of 250000–3000000 of PBMCs or 100–30000 of purified B cells per well in duplicate. After incubation for 16 h at 37°C, 5% CO_2_, the cells were thoroughly removed with washing buffer (0.05% Tween 20 in PBS). Isotype-specific ASCs were detected using IgG- or IgM-specific biotinylated rabbit antibodies (R&D Systems, Cat. No. SELB002, SELB003) or IgA-specific biotinylated goat antibodies (SouthernBiotech, Cat. No. 2052-08).

After five sequential washes with 0.05% Tween-20 / PBS, streptavidin alkaline phosphatase conjugate (R&D Systems, Cat. No. SEL002) was added at a 1:60 dilution and the plates were further incubated for 2 hours at room temperature. After several washes, the colorimetric reaction was developed by the addition of Substrate Reagent from B Cell ELISpot Development Module (R&B Systems, Cat. No. SEL002) until clear distinct spots appeared. The reaction was stopped by rinsing the plate with tap water. ELISpot images were acquired using the CTL ImmunoSpot® analyzer (CTL). Spots were counted using ImmunoSpot® software. Wells coated with an irrelevant protein Bet v 1 served as negative controls.

### Pseudotyped virus neutralization (pVNT) assay

For titration of the neutralizing activity of plasma samples, lentiviral particles pseudotyped with the SARS-CoV-2 S-protein of the ancestral Wuhan strain, or B1.351 VOC were used. Lentiviral particles were produced as follows. HEK293T cells were transfected with plasmids psPax2 (kind gift from Dr. Didier Trono), pLV-eGFP (was a gift from Pantelis Tsoulfas, (Addgene, Cat. No. 36083)), and the pCAGGS-SΔ19 plasmid encoding wild-type or B1.351 S protein (see below). Transfection was performed by the calcium phosphate method (Kutner, Zhang,, Reiser 2009); 72 hours after transfection, the supernatants were filtered through a 0.45 μm filter, concentrated 20-fold on Amicon® Ultra-15 ultrafiltration cells with a 100 kDa cutoff (Merck, Cat. No. UFC910008). Concentrated supernatant was further centrifuged at 20,000 g, 8°C for 90 min. The pellet was resuspended in Opti-MEM medium. Then the viral particles were immediately used in neutralization tests or stored at -70°C for no more than a month. Viral yield was quantified using titration on HEK293T-hACE2 cells.

Prior to analysis, plasma samples were heated for 30 min at 56°C to inactivate complement. After that, serial two-fold plasma dilutions ranging 1:10 - 1:1280 were prepared in a 96-well plate and 20,000 lentiviral particles were added in an equal volume of Opti-Mem supplemented with 2.5% of heat-inactivated FBS. Plasma and viral particles were co-incubated for 30 min at 37°C, and added to HEK293T-hACE2 cells. 72 hours following transduction, the percentage of transduced cells was measured in the cultures using flow cytometry. Half-maximal inhibitory dilution (ID50) was determined by non-linear regression as the serum dilution that neutralized 50% of the pseudotyped lentivirus.

### Cloning of the B.1.351 Spike variant

The construct pCAGGS-SΔ19 carrying a codon-optimized cassette encoding a SARS-CoV-2 Spike protein (reference Wuhan-Hu-1 isolate) lacking 19 C-terminal residues, which has been shown to boost the viral titers (Johnson et al., 2020), has been described (Gorchakov et al., 2021 in press).To obtain pCAGGS-SΔ19_B.1.351, sets of complementary mutagenic primers (27 nt each) centered at the desired site were used to sequentially introduce the individual mutations (L18F, D80A, D215G, del241-243, R246I, K417N, E484K, N501Y, D614G, A701V) into the coding sequence of SΔ19. Sequence identity of the resulting plasmid was confirmed by Sanger sequencing.

### Statistical analysis

Statistical analysis was performed using Graph Pad Prism (version 8.4.3 GraphPad Software, La Jolla California). The Kruskal–Wallis H test was used for comparison between multiple groups. P < 0.05 was considered statistically significant. Calculation of 95% confidence intervals (CI) was based on the t-distribution of the log-transformed titers, then back-transformed to the original scale. The correlation between two groups was determined by Spearman rank test. A normalized non-linear regression was performed using GraphPad Prism software (Sigmoidal, 4PL). Heatmap generation and principal component analysis were performed with Clustvis (Metsalu, Vilo 2015) using normalized data. Data are presented as median ± IQR. Asterisks indicate significant difference between groups determined using the Kruskal–Wallis test, *P < 0.05, **P < 0.01, ***P < 0.001, ****P < 0.0001, ns = not significant.

## Data Availability

All data produced in the present work are contained in the manuscript

## Acknowledgments

The authors thank all the individuals in the study for the kind donation of both their time and biological material. We thank Gaukhar Yusubalieva, Vladimir Baklaushev, Yuri Lebedin and Rudolf Valenta for their kind help with experiments and for providing reagents. Fig. 1a and Supplementary Fig. 1 were created with the help of BioRender.com. This work was supported by the Russian Science Foundation (Project 21-15-00331) and the Russian Fund for Basic Research (20-04-60527).

## Additional files

Supplementary file

Transparent reporting form

---

**Supplementary table 1.** Participant characteristics.

| Recipient ID | Sex | Age range | COVID-19 symptoms<sup>§</sup> | Vaccination symptoms |  |
| --- | --- | --- | --- | --- | --- |
|  |  |  |  | 1st dose | 2nd dose |
| 1 | M | 61-65 | -70 to -61<sup>§§</sup> | 37.8°C | 38.7°C |
| 2 | M | 66-70 | -53 to -46<sup>§§</sup> | injection site reaction | w/o |
| 3 | F | 61-65 | w/o | w/o | w/o |
| 4 | F | 56-60 | w/o | w/o | w/o |
| 5 | M | 56-60 | w/o | w/o | w/o |
| 6 | M | 66-70 | w/o | headache | w/o |
| 7 | F | 66-70 | w/o | w/o | w/o |
| 8 | F | 51-55 | w/o | w/o | w/o |
| 9 | M | 66-70 | w/o | w/o | w/o |
| 10 | F | 41-45 | w/o | w/o | w/o |
| 11 | F | 61-65 | w/o | w/o | w/o |
| 12 | F | 51-55 | w/o | w/o | w/o |
| 13 | F | 21-25 | w/o | w/o | w/o |
| 14 | F | 46-50 | w/o | w/o | w/o |
| 15 | F | 41-45 | w/o | w/o | w/o |
| 16 | F | 51-55 | w/o | w/o | injection site reaction |
| 17 | M | 41-45 | -86 to -78<sup>§§</sup> | 37.8°C | 38.7°C |
| 18 | M | 61-65 | -120 to -110<sup>§§</sup> | 37.8°C | w/o |
| 19 | F | 66-70 | w/o | w/o | w/o |
| 20 | F | 61-65 | w/o | w/o | w/o |
| 21 | F | 51-55 | -88 to -71<sup>§§</sup> | w/o | w/o |
| 22 | F | 51-55 | w/o | 37.5°C | 37.5°C |

<sup>§</sup> mild COVID-19 symptoms (cough, sore throat, headache, runny nose, and muscle ache) were observed on the indicated days.
<sup>§§</sup> PCR-confirmed COVID-19, two months after vaccination

**Supplementary Fig. 1.**
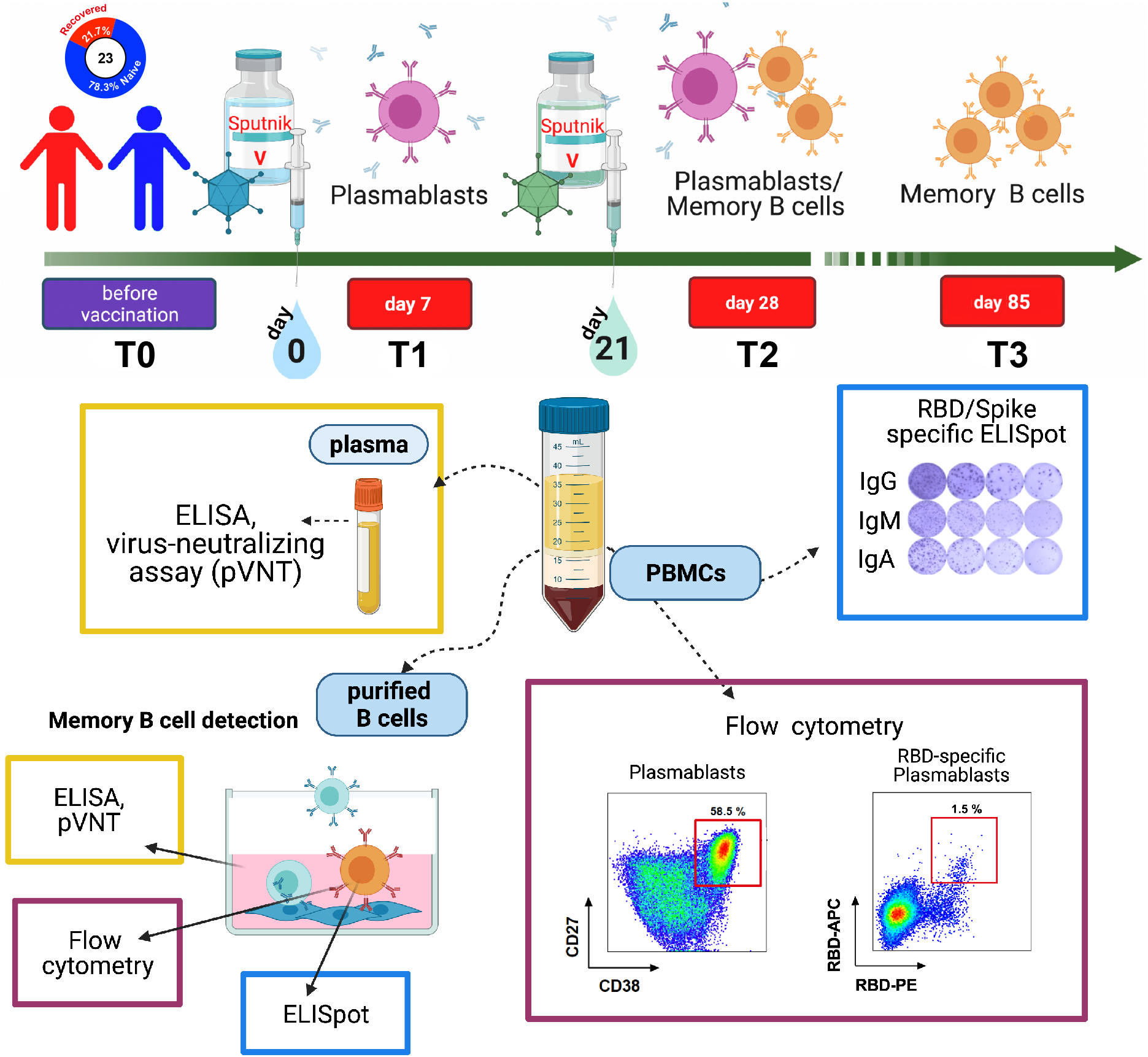
Study design.

**Supplementary Figure 2.**
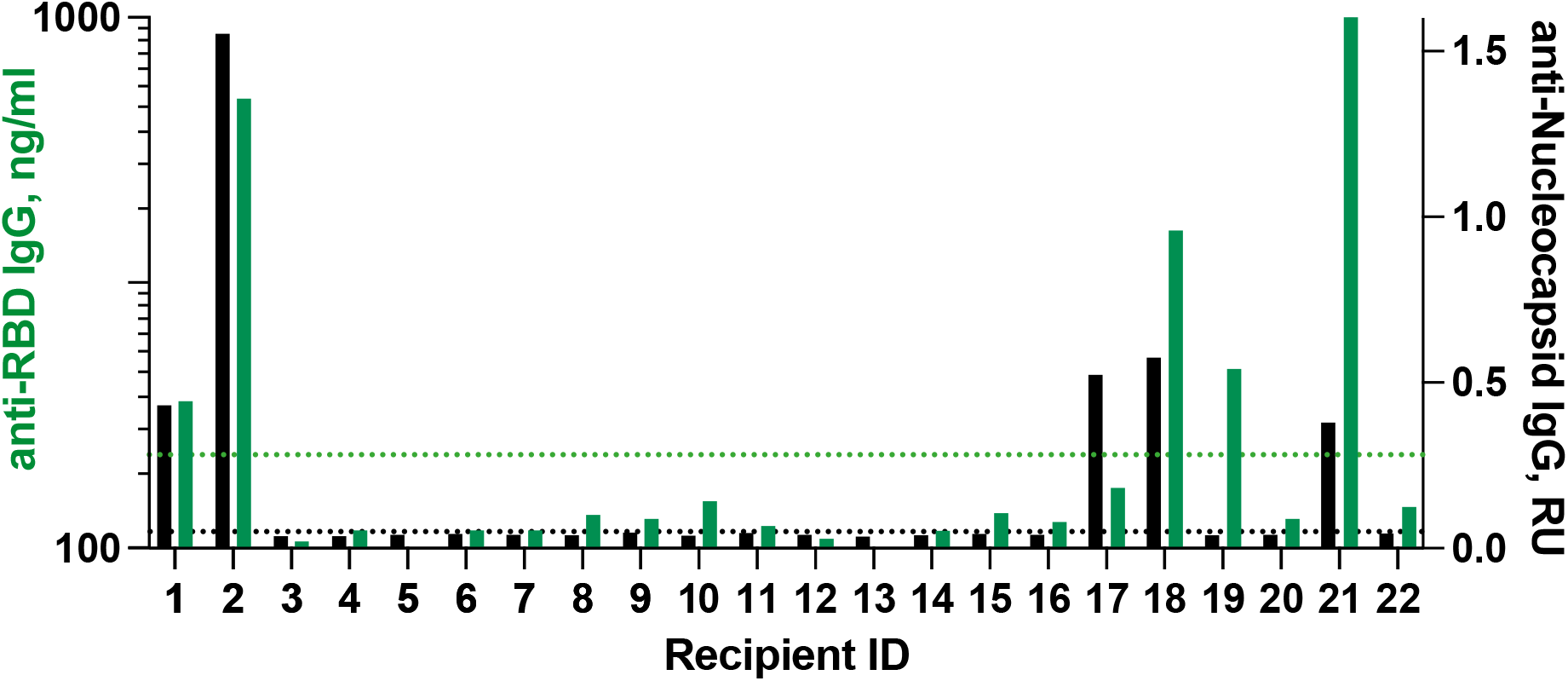
IgG antibodies against the receptor-binding domain (RBD) (green bars), and nucleocapsid (N) (black bars) of SARS-CoV-2 at T0 time point just before vaccination.

**Supplementary Figure 3.**
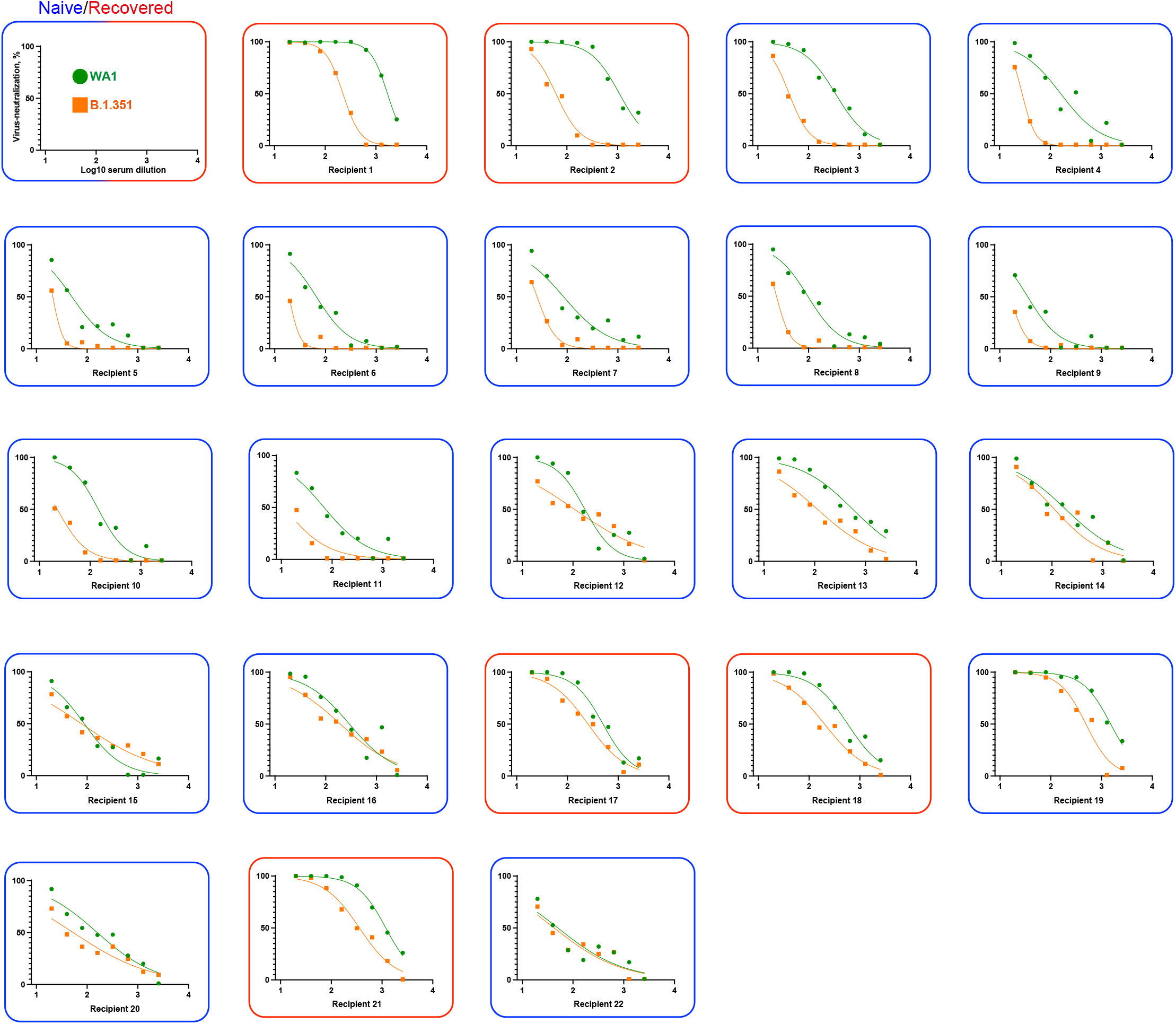
Neutralization curves of vaccine-induced sera against pseudotyped virus expressing SARS-CoV-2 WA1 or B.1.351 variant Spike protein. Neutralization activities of sera from naïve (n=17, blue frames) and recovered (n=5, red frames) individuals were measured at T3 time point.

## References

Abayasingam A, Balachandran H, Agapiou D, Hammoud M, Rodrigo C, Keoshkerian E, Li H, Brasher NA, Christ D, Rouet R, Burnet D, Grubor-Bauk B, Rawlinson W, Turville S, Aggarwal A, Stella AO, Fichter C, Brilot F, Mina M, Post JJ, Hudson B, Gilroy N, Dwyer D, Sasson SC, Tea F, Pilli D, Kelleher A, Tedla N, Lloyd AR, Martinello M, Bull RA; COSIN Study Group. Long-term persistence of RBD+ memory B cells encoding neutralizing antibodies in SARSCoV-2 infection. Cell Rep Med. 2021; 2(4):100228. doi: 10.1016/j.xcrm.2021.100228.

Abdool Karim SS, de Oliveira T. New SARS-CoV-2 Variants - Clinical, Public Health, and Vaccine Implications. N Engl J Med. 2021; 384(19):1866–1868. doi: 10.1056/NEJMc2100362.

Anderson EJ, Rouphael NG, Widge AT, Jackson LA, Roberts PC, Makhene M, Chappell JD, Denison MR, Stevens LJ, Pruijssers AJ, McDermott AB, Flach B, Lin BC, Doria-Rose NA, O’Dell S, Schmidt SD, Corbett KS, Swanson PA 2nd, Padilla M, Neuzil KM, Bennett H, Leav B, Makowski M, Albert J, Cross K, Edara VV, Floyd K, Suthar MS, Martinez DR, Baric R, Buchanan W, Luke CJ, Phadke VK, Rostad CA, Ledgerwood JE, Graham BS, Beigel JH; mRNA-1273 Study Group. Safety and Immunogenicity of SARS-CoV-2 mRNA-1273 Vaccine in Older Adults. N Engl J Med. 2020; 383(25):2427–2438. doi: 10.1056/NEJMoa2028436.

Appanna R, Kg S, Xu MH, Toh YX, Velumani S, Carbajo D, Lee CY, Zuest R, Balakrishnan T, Xu W, Lee B, Poidinger M, Zolezzi F, Leo YS, Thein TL, Wang CI, Fink K. Plasmablasts During Acute Dengue Infection Represent a Small Subset of a Broader Virus-specific Memory B Cell Pool. EBioMedicine. 2016; 12:178–188. doi: 10.1016/j.ebiom.2016.09.003.

Bos R, Rutten L, van der Lubbe JEM, Bakkers MJG, Hardenberg G, Wegmann F, Zuijdgeest D, de Wilde AH, Koornneef A, Verwilligen A, van Manen D, Kwaks T, Vogels R, Dalebout TJ, Myeni SK, Kikkert M, Snijder EJ, Li Z, Barouch DH, Vellinga J, Langedijk JPM, Zahn RC, Custers J, Schuitemaker H. Ad26 vector-based COVID-19 vaccine encoding a prefusionstabilized SARS-CoV-2 Spike immunogen induces potent humoral and cellular immuneresponses. NPJ Vaccines. 2020; 5:91. doi: 10.1038/s41541-020-00243-x.

Bucci EM, Berkhof J, Gillibert A, Gopalakrishna G, Calogero RA, Bouter LM, Andreev K, Naudet F, Vlassov V. Data discrepancies and substandard reporting of interim data of Sputnik V phase 3 trial. Lancet. 2021; 397(10288):1881–1883. doi: 10.1016/S0140-6736(21)00899-0.

Byazrova M, Yusubalieva G, Spiridonova A, Efimov G, Mazurov D, Baranov K, Baklaushev V, Filatov A. Pattern of circulating SARS-CoV-2-specific antibody-secreting and memory B-cell generation in patients with acute COVID-19. Clin Transl Immunology. 2021; 10(2):e1245. doi: 10.1002/cti2.1245.

Dan JM, Mateus J, Kato Y, Hastie KM, Yu ED, Faliti CE, Grifoni A, Ramirez SI, Haupt S, Frazier A, Nakao C, Rayaprolu V, Rawlings SA, Peters B, Krammer F, Simon V, Saphire EO, Smith DM, Weiskopf D, Sette A, Crotty S. Immunological memory to SARS-CoV-2 assessed for up to 8 months after infection. Science. 2021; 371(6529):eabf4063. doi: 10.1126/science.abf4063.

Davies NG, Abbott S, Barnard RC, Jarvis CI, Kucharski AJ, Munday JD, Pearson CAB, Russell TW, Tully DC, Washburne AD, Wenseleers T, Gimma A, Waites W, Wong KLM, van Zandvoort K, Silverman JD; CMMID COVID-19 Working Group; COVID-19 Genomics UK (COG-UK) Consortium, Diaz-Ordaz K, Keogh R, Eggo RM, Funk S, Jit M, Atkins KE, Edmunds WJ. Estimated transmissibility and impact of SARS-CoV-2 lineage B.1.1.7 in England. Science. 2021; 372(6538):eabg3055. doi: 10.1126/science.abg3055.

Dejnirattisai W, Zhou D, Ginn HM, Duyvesteyn HME, Supasa P, Case JB, Zhao Y, Walter TS, Mentzer AJ, Liu C, Wang B, Paesen GC, Slon-Campos J, Lopez-Camacho C, Kafai NM, Bailey AL, Chen RE, Ying B, Thompson C, Bolton J, Fyfe A, Gupta S, Tan TK, Gilbert-Jaramillo J, James W, Knight M, Carroll MW, Skelly D, Dold C, Peng Y, Levin R, Dong T, Pollard AJ, Knight JC, Klenerman P, Temperton N, Hall DR, Williams MA, Paterson NG, Bertram FKR, Siebert CA, Clare DK, Howe A, Radecke J, Song Y, Townsend AR, Huang KA, Fry EE, Mongkolsapaya J, Diamond MS, Ren J, Stuart DI, Screaton GR. The antigenic anatomy of SARS-CoV-2 receptor binding domain. Cell. 2021; 184(8):2183-2200.e22. doi: 10.1016/j.cell.2021.02.032.

Dejnirattisai W, Zhou D, Supasa P, Liu C, Mentzer AJ, Ginn HM, Zhao Y, Duyvesteyn HME, Tuekprakhon A, Nutalai R, Wang B, Lopez-Camacho C, Slon-Campos J, Walter TS, Skelly D, Costa Clemens SA, Naveca FG, Nascimento V, Nascimento F, Fernandes da Costa C, Resende PC, Pauvolid-Correa A, Siqueira MM, Dold C, Levin R, Dong T, Pollard AJ, Knight JC, Crook D, Lambe T, Clutterbuck E, Bibi S, Flaxman A, Bittaye M, Belij-Rammerstorfer S, Gilbert SC, Carroll MW, Klenerman P, Barnes E, Dunachie SJ, Paterson NG, Williams MA, Hall DR, Hulswit RJG, Bowden TA, Fry EE, Mongkolsapaya J, Ren J, Stuart DI, Screaton GR. Antibody evasion by the P.1 strain of SARS-CoV-2. Cell. 2021; 184(11):2939-2954.e9. doi: 10.1016/j.cell.2021.03.055.

Ding BB, Bi E, Chen H, Yu JJ, Ye BH. IL-21 and CD40L synergistically promote plasma cell differentiation through upregulation of Blimp-1 in human B cells. J Immunol. 2013; 190(4):1827–36. doi: 10.4049/jimmunol.1201678.

Ebinger JE, Fert-Bober J, Printsev I, Wu M, Sun N, Prostko JC, Frias EC, Stewart JL, Van Eyk JE, Braun JG, Cheng S, Sobhani K. Antibody responses to the BNT162b2 mRNA vaccine in individuals previously infected with SARS-CoV-2. Nat Med. 2021; 27(6):981–984. doi: 10.1038/s41591-021-01325-6.

Folegatti PM, Ewer KJ, Aley PK, Angus B, Becker S, Belij-Rammerstorfer S, Bellamy D, Bibi S, Bittaye M, Clutterbuck EA, Dold C, Faust SN, Finn A, Flaxman AL, Hallis B, Heath P, Jenkin D, Lazarus R, Makinson R, Minassian AM, Pollock KM, Ramasamy M, Robinson H, Snape M, Tarrant R, Voysey M, Green C, Douglas AD, Hill AVS, Lambe T, Gilbert SC, Pollard AJ; Oxford COVID Vaccine Trial Group. Safety and immunogenicity of the ChAdOx1 nCoV-19 vaccine against SARS-CoV-2: a preliminary report of a phase 1/2, single-blind, randomized controlled trial. Lancet. 2020; 396(10249):467–478. doi: 10.1016/S0140-6736(20)31604-4.

Garcia-Beltran WF, Lam EC, St Denis K, Nitido AD, Garcia ZH, Hauser BM, Feldman J, Pavlovic MN, Gregory DJ, Poznansky MC, Sigal A, Schmidt AG, Iafrate AJ, Naranbhai V, Balazs AB. Multiple SARS-CoV-2 variants escape neutralization by vaccine-induced humoral immunity. Cell. 2021; 184(9):2372-2383.e9. doi: 10.1016/j.cell.2021.03.013.

Goel RR, Apostolidis SA, Painter MM, Mathew D, Pattekar A, Kuthuru O, Gouma S, Hicks P, Meng W, Rosenfeld AM, Dysinger S, Lundgreen KA, Kuri-Cervantes L, Adamski S, Hicks A, Korte S, Oldridge DA, Baxter AE, Giles JR, Weirick ME, McAllister CM, Dougherty J, Long S, D’Andrea K, Hamilton JT, Betts MR, Luning Prak ET, Bates P, Hensley SE, Greenplate AR, Wherry EJ. Distinct antibody and memory B cell responses in SARS-CoV-2 naive and recovered individuals following mRNA vaccination. Sci Immunol. 2021; 6(58):eabi6950. doi: 10.1126/sciimmunol.abi6950.

Gushchin VA, Dolzhikova IV, Shchetinin AM, Odintsova AS, Siniavin AE, Nikiforova MA, Pochtovyi AA, Shidlovskaya EV, Kuznetsova NA, Burgasova OA, Kolobukhina LV, Iliukhina AA, Kovyrshina AV, Botikov AG, Kuzina AV, Grousova DM, Tukhvatulin AI, Shcheblyakov DV, Zubkova OV, Karpova OV, Voronina OL, Ryzhova NN, Aksenova EI, Kunda MS, Lioznov DA, Danilenko DM, Komissarov AB, Tkachuck AP, Logunov DY, Gintsburg AL. Neutralizing Activity of Sera from Sputnik V-Vaccinated People against Variants of Concern (VOC: B.1.1.7, B.1.351, P.1, B.1.617.2, B.1.617.3) and Moscow Endemic SARS-CoV-2 Variants. Vaccines (Basel). 2021; 9(7):779. doi: 10.3390/vaccines9070779.

Hartley GE, Edwards ESJ, Aui PM, Varese N, Stojanovic S, McMahon J, Peleg AY, Boo I, Drummer HE, Hogarth PM, O’Hehir RE, van Zelm MC. Rapid generation of durable B cell memory to SARS-CoV-2 spike and nucleocapsid proteins in COVID-19 and convalescence. Sci Immunol. 2020; 5(54):eabf8891. doi: 10.1126/sciimmunol.abf8891.

Ikegame S, Siddiquey MNA, Hung CT, Haas G, Brambilla L, Oguntuyo KY, Kowdle S, Chiu HP, Stevens CS, Vilardo AE, Edelstein A, Perandones C, Kamil JP, Lee B. Neutralizing activity of Sputnik V vaccine sera against SARS-CoV-2 variants. Nat Commun. 2021; 12(1):4598. doi: 10.1038/s41467-021-24909-9.

Johnson MC, Lyddon TD, Suarez R, Salcedo B, LePique M, Graham M, Ricana C, Robinson C, Ritter DG. Optimized Pseudotyping Conditions for the SARS-COV-2 Spike Glycoprotein. J Virol. 2020; 94(21):e01062–20. doi: 10.1128/JVI.01062-20.

Khvastunova AN, Kuznetsova SA, Al-Radi LS, Vylegzhanina AV, Zakirova AO, Fedyanina OS, Filatov AV, Vorobjev IA, Ataullakhanov F. Anti-CD antibody microarray for human leukocyte morphology examination allows analyzing rare cell populations and suggesting preliminary diagnosis in leukemia. Sci Rep. 2015; 5:12573. doi: 10.1038/srep12573.

Krammer F, Srivastava K, Alshammary H, Amoako AA, Awawda MH, Beach KF, Bermudez-Gonzalez MC, Bielak DA, Carreno JM, Chernet RL, Eaker LQ, Ferreri ED, Floda DL, Gleason CR, Hamburger JZ, Jiang K, Kleiner G, Jurczyszak D, Matthews JC, Mendez WA, Nabeel I, Mulder LCF, Raskin AJ, Russo KT, Salimbangon AT, Saksena M, Shin AS, Singh G, Sominsky LA, Stadlbauer D, Wajnberg A, Simon V. Antibody Responses in Seropositive Persons after a Single Dose of SARS-CoV-2 mRNA Vaccine. N Engl J Med. 2021; 384(14):1372–1374. doi: 10.1056/NEJMc2101667.

Kuri-Cervantes L, Pampena MB, Meng W, Rosenfeld AM, Ittner CAG, Weisman AR, Agyekum RS, Mathew D, Baxter AE, Vella LA, Kuthuru O, Apostolidis SA, Bershaw L, Dougherty J, Greenplate AR, Pattekar A, Kim J, Han N, Gouma S, Weirick ME, Arevalo CP, Bolton MJ, Goodwin EC, Anderson EM, Hensley SE, Jones TK, Mangalmurti NS, Luning Prak ET, Wherry EJ, Meyer NJ, Betts MR. Comprehensive mapping of immune perturbations associated with severe COVID-19. Sci Immunol. 2020; 5(49):eabd7114. doi: 10.1126/sciimmunol.abd7114.

Kustin T, Harel N, Finkel U, Perchik S, Harari S, Tahor M, Caspi I, Levy R, Leshchinsky M, Ken Dror S, Bergerzon G, Gadban H, Gadban F, Eliassian E, Shimron O, Saleh L, Ben-Zvi H, Keren Taraday E, Amichay D, Ben-Dor A, Sagas D, Strauss M, Shemer Avni Y, Huppert A, Kepten E, Balicer RD, Netzer D, Ben-Shachar S, Stern A. Evidence for increased breakthrough rates of SARS-CoV-2 variants of concern in BNT162b2-mRNA-vaccinated individuals. Nat Med. 2021; 27(8):1379–1384. doi: 10.1038/s41591-021-01413-7.

Kutner RH, Zhang XY, Reiser J. Production, concentration and titration of pseudotyped HIV-1-based lentiviral vectors. Nat Protoc. 2009; 4(4):495–505. doi: 10.1038/nprot.2009.22.

Liu Y, Liu J, Xia H, Zhang X, Fontes-Garfias CR, Swanson KA, Cai H, Sarkar R, Chen W, Cutler M, Cooper D, Weaver SC, Muik A, Sahin U, Jansen KU, Xie X, Dormitzer PR, Shi PY. Neutralizing Activity of BNT162b2-Elicited Serum. N Engl J Med. 2021; 384(15):1466–1468. doi: 10.1056/NEJMc2102017.

Logunov DY, Dolzhikova IV, Shcheblyakov DV, Tukhvatulin AI, Zubkova OV, Dzharullaeva AS, Kovyrshina AV, Lubenets NL, Grousova DM, Erokhova AS, Botikov AG, Izhaeva FM, Popova O, Ozharovskaya TA, Esmagambetov IB, Favorskaya IA, Zrelkin DI, Voronina DV, Shcherbinin DN, Semikhin AS, Simakova YV, Tokarskaya EA, Egorova DA, Shmarov MM, Nikitenko NA, Gushchin VA, Smolyarchuk EA, Zyryanov SK, Borisevich SV, Naroditsky BS, Gintsburg AL; Gam-COVID-Vac Vaccine Trial Group. Safety and efficacy of an rAd26 and rAd5 vector-based heterologous prime-boost COVID-19 vaccine: an interim analysis of a randomised controlled phase 3 trial in Russia. Lancet. 2021; 397(10275):671–681. doi: 10.1016/S0140-6736(21)00234-8.

Logunov DY, Dolzhikova IV, Zubkova OV, Tukhvatulin AI, Shcheblyakov DV, Dzharullaeva AS, Grousova DM, Erokhova AS, Kovyrshina AV, Botikov AG, Izhaeva FM, Popova O, Ozharovskaya TA, Esmagambetov IB, Favorskaya IA, Zrelkin DI, Voronina DV, Shcherbinin DN, Semikhin AS, Simakova YV, Tokarskaya EA, Lubenets NL, Egorova DA, Shmarov MM, Nikitenko NA, Morozova LF, Smolyarchuk EA, Kryukov EV, Babira VF, Borisevich SV, Naroditsky BS, Gintsburg AL. Safety and immunogenicity of an rAd26 and rAd5 vector-based heterologous prime-boost COVID-19 vaccine in two formulations: two open, non-randomised phase 1/2 studies from Russia. Lancet. 2020; 396(10255):887–897. doi: 10.1016/S0140-6736(20)31866-3.

Metsalu T, Vilo J. ClustVis: a web tool for visualizing clustering of multivariate data using Principal Component Analysis and heatmap. Nucleic Acids Res. 2015; 43(W1):W566-70. doi: 10.1093/nar/gkv468.

Naveca FG, Nascimento V, de Souza VC, Corado AL, Nascimento F, Silva G, Costa A, Duarte D, Pessoa K, Mejia M, Brandao MJ, Jesus M, Goncalves L, da Costa CF, Sampaio V, Barros D, Silva M, Mattos T, Pontes G, Abdalla L, Santos JH, Arantes I, Dezordi FZ, Siqueira MM, Wallau GL, Resende PC, Delatorre E, Graf T, Bello G. COVID-19 in Amazonas, Brazil, was driven by the persistence of endemic lineages and P.1 emergence. Nat Med. 2021; 27(7):1230–1238. doi: 10.1038/s41591-021-01378-7.

Ortloff A, Harsch IA. Outbreak of infections with the COVID-19 virus mutation B.1.351 about four weeks after the second successful vaccination with BNT162b2. GMS Hyg Infect Control. 2021; 16:Doc16. doi: 10.3205/dgkh000387.

Rossi AH, Ojeda DS, Varese A, Sanchez L, Gonzalez Lopez Ledesma MM, Mazzitelli I, Alvarez Julia A, Oviedo Rouco S, Pallares HM, Costa Navarro GS, Rasetto NB, Garcia CI, Wenker SD, Ramis LY, Bialer MG, de Leone MJ, Hernando CE, Sosa S, Bianchimano L, Rios AS, Treffinger Cienfuegos MS, Caramelo JJ, Longueira Y, Laufer N, Alvarez DE, Carradori J, Pedrozza D, Rima A, Echegoyen C, Ercole R, Gelpi P, Marchetti S, Zubieta M, Docena G, Kreplak N, Yanovsky M, Geffner J, Pifano M, Gamarnik AV. Sputnik V vaccine elicits seroconversion and neutralizing capacity to SARS-CoV-2 after a single dose. Cell Rep Med. 2021; 2(8):100359. doi: 10.1016/j.xcrm.2021.100359.

Sanz I, Wei C, Jenks SA, Cashman KS, Tipton C, Woodruff MC, Hom J, Lee FE. Challenges and Opportunities for Consistent Classification of Human B Cell and Plasma Cell Populations. Front Immunol. 2019; 10:2458. doi: 10.3389/fimmu.2019.02458.

Sasikala M, Shashidhar J, Deepika G, Ravikanth V, Krishna VV, Sadhana Y, Pragathi K, Reddy DN. Immunological memory and neutralizing activity to a single dose of COVID-19 vaccine in previously infected individuals. Int J Infect Dis. 2021; 108:183–186. doi: 10.1016/j.ijid.2021.05.034.

Scheid JF, Barnes CO, Eraslan B, Hudak A, Keeffe JR, Cosimi LA, Brown EM, Muecksch F, Weisblum Y, Zhang S, Delorey T, Woolley AE, Ghantous F, Park SM, Phillips D, Tusi B, Huey-Tubman KE, Cohen AA, Gnanapragasam PNP, Rzasa K, Hatziioanno T, Durney MA, Gu X, Tada T, Landau NR, West AP Jr, Rozenblatt-Rosen O, Seaman MS, Baden LR, Graham DB, Deguine J, Bieniasz PD, Regev A, Hung D, Bjorkman PJ, Xavier RJ. B cell genomics behind cross-neutralization of SARS-CoV-2 variants and SARS-CoV. Cell. 2021; 184(12):3205-3221.e24. doi: 10.1016/j.cell.2021.04.032.

Shomuradova AS, Vagida MS, Sheetikov SA, Zornikova KV, Kiryukhin D, Titov A, Peshkova IO, Khmelevskaya A, Dianov DV, Malasheva M, Shmelev A, Serdyuk Y, Bagaev DV, Pivnyuk A, Shcherbinin DS, Maleeva AV, Shakirova NT, Pilunov A, Malko DB, Khamaganova EG, Biderman B, Ivanov A, Shugay M, Efimov GA. SARS-CoV-2 Epitopes Are Recognized by a Public and Diverse Repertoire of Human T Cell Receptors. Immunity. 2020; 53(6):1245-1257.e5. doi: 10.1016/j.immuni.2020.11.004.

Stamatatos L, Czartoski J, Wan YH, Homad LJ, Rubin V, Glantz H, Neradilek M, Seydoux E, Jennewein MF, MacCamy AJ, Feng J, Mize G, De Rosa SC, Finzi A, Lemos MP, Cohen KW, Moodie Z, McElrath MJ, McGuire AT. mRNA vaccination boosts cross-variant neutralizing antibodies elicited by SARS-CoV-2 infection. Science. 2021: eabg9175. doi: 10.1126/science.abg9175.

Tegally H, Wilkinson E, Giovanetti M, Iranzadeh A, Fonseca V, Giandhari J, Doolabh D, Pillay S, San EJ, Msomi N, Mlisana K, von Gottberg A, Walaza S, Allam M, Ismail A, Mohale T, Glass AJ, Engelbrecht S, Van Zyl G, Preiser W, Petruccione F, Sigal A, Hardie D, Marais G, Hsiao NY, Korsman S, Davies MA, Tyers L, Mudau I, York D, Maslo C, Goedhals D, Abrahams S, Laguda-Akingba O, Alisoltani-Dehkordi A, Godzik A, Wibmer CK, Sewell BT, Lourenco J, Alcantara LCJ, Kosakovsky Pond SL, Weaver S, Martin D, Lessells RJ, Bhiman JN, Williamson C, de Oliveira T. Detection of a SARS-CoV-2 variant of concern in South Africa. Nature. 2021; 592(7854):438–443. doi: 10.1038/s41586-021-03402-9.

Wang P, Nair MS, Liu L, Iketani S, Luo Y, Guo Y, Wang M, Yu J, Zhang B, Kwong PD, Graham BS, Mascola JR, Chang JY, Yin MT, Sobieszczyk M, Kyratsous CA, Shapiro L, Sheng Z, Huang Y, Ho DD. Antibody resistance of SARS-CoV-2 variants B.1.351 and B.1.1.7. Nature. 2021; 593(7857):130–135. doi: 10.1038/s41586-021-03398-2.

Wang Z, Muecksch F, Schaefer-Babajew D, Finkin S, Viant C, Gaebler C, Hoffmann HH, Barnes CO, Cipolla M, Ramos V, Oliveira TY, Cho A, Schmidt F, Da Silva J, Bednarski E, Aguado L, Yee J, Daga M, Turroja M, Millard KG, Jankovic M, Gazumyan A, Zhao Z, Rice CM, Bieniasz PD, Caskey M, Hatziioannou T, Nussenzweig MC. Naturally enhanced neutralizing breadth against SARS-CoV-2 one year after infection. Nature. 2021; 595(7867):426–431. doi: 10.1038/s41586-021-03696-9.

Wang Z, Schmidt F, Weisblum Y, Muecksch F, Barnes CO, Finkin S, Schaefer-Babajew D, Cipolla M, Gaebler C, Lieberman JA, Oliveira TY, Yang Z, Abernathy ME, Huey-Tubman KE, Hurley A, Turroja M, West KA, Gordon K, Millard KG, Ramos V, Da Silva J, Xu J, Colbert RA, Patel R, Dizon J, Unson-O’Brien C, Shimeliovich I, Gazumyan A, Caskey M, Bjorkman PJ, Casellas R, Hatziioannou T, Bieniasz PD, Nussenzweig MC. mRNA vaccine-elicited antibodies to SARS-CoV-2 and circulating variants. Nature. 2021; 592(7855):616–622. doi: 10.1038/s41586-021-03324-6.

Wibmer CK, Ayres F, Hermanus T, Madzivhandila M, Kgagudi P, Oosthuysen B, Lambson BE, de Oliveira T, Vermeulen M, van der Berg K, Rossouw T, Boswell M, Ueckermann V, Meiring S, von Gottberg A, Cohen C, Morris L, Bhiman JN, Moore PL. SARS-CoV-2 501Y.V2 escapes neutralization by South African COVID-19 donor plasma. Nat Med. 2021; 27(4):622–625. doi: 10.1038/s41591-021-01285-x.

Woodruff MC, Ramonell RP, Nguyen DC, Cashman KS, Saini AS, Haddad NS, Ley AM, Kyu S, Howell JC, Ozturk T, Lee S, Suryadevara N, Case JB, Bugrovsky R, Chen W, Estrada J, Morrison-Porter A, Derrico A, Anam FA, Sharma M, Wu HM, L. SN, Jenks SA, Tipton CM, Staitieh B, Daiss JL, Ghosn E, Diamond MS, Carnahan RH, Crowe JE Jr, Hu WT, Lee FE, Sanz I. Extrafollicular B cell responses correlate with neutralizing antibodies and morbidity in COVID-19. Nat Immunol. 2020; 21(12):1506–1516. doi: 10.1038/s41590-020-00814-z.

Wrammert J, Smith K, Miller J, Langley WA, Kokko K, Larsen C, Zheng NY, Mays I, Garman L, Helms C, James J, Air GM, Capra JD, Ahmed R, Wilson PC. Rapid cloning of high-affinity human monoclonal antibodies against influenza virus. Nature. 2008; 453(7195):667–71. doi: 10.1038/nature06890.

Wu K, Werner AP, Koch M, Choi A, Narayanan E, Stewart-Jones GBE, Colpitts T, Bennett H, Boyoglu-Barnum S, Shi W, Moliva JI, Sullivan NJ, Graham BS, Carfi A, Corbett KS, Seder RA, Edwards DK. Serum Neutralizing Activity Elicited by mRNA-1273 Vaccine. N Engl J Med. 2021; 384(15):1468–1470. doi: 10.1056/NEJMc2102179.

Yang Y, Du L. SARS-CoV-2 spike protein: a key target for eliciting persistent neutralizing antibodies. Signal Transduct Target Ther. 2021; 6(1):95. doi: 10.1038/s41392-021-00523-5.

Yao L, Wang GL, Shen Y, Wang ZY, Zhan BD, Duan LJ, Lu B, Shi C, Gao YM, Peng HH, Wang GQ, Wang DM, Jiang MD, Cao GP, Ma MJ. Persistence of Antibody and Cellular Immune Responses in Coronavirus Disease 2019 Patients Over Nine Months After Infection. J Infect Dis. 2021; 224(4):586–594. doi: 10.1093/infdis/jiab255.

Zhou D, Dejnirattisai W, Supasa P, Liu C, Mentzer AJ, Ginn HM, Zhao Y, Duyvesteyn HME, Tuekprakhon A, Nutalai R, Wang B, Paesen GC, Lopez-Camacho C, Slon-Campos J, Hallis B, Coombes N, Bewley K, Charlton S, Walter TS, Skelly D, Lumley SF, Dold C, Levin R, Dong T, Pollard AJ, Knight JC, Crook D, Lambe T, Clutterbuck E, Bibi S, Flaxman A, Bittaye M, Belij-Rammerstorfer S, Gilbert S, James W, Carroll MW, Klenerman P, Barnes E, Dunachie SJ, Fry EE, Mongkolsapaya J, Ren J, Stuart DI, Screaton GR. Evidence of escape of SARS-CoV-2 variant B.1.351 from natural and vaccine-induced sera. Cell. 2021; 184(9):2348-2361.e6. doi: 10.1016/j.cell.2021.02.037.

Zhu FC, Guan XH, Li YH, Huang JY, Jiang T, Hou LH, Li JX, Yang BF, Wang L, Wang WJ, Wu SP, Wang Z, Wu XH, Xu JJ, Zhang Z, Jia SY, Wang BS, Hu Y, Liu JJ, Zhang J, Qian XA, Li Q, Pan HX, Jiang HD, Deng P, Gou JB, Wang XW, Wang XH, Chen W. Immunogenicity and safety of a recombinant adenovirus type-5-vectored COVID-19 vaccine in healthy adults aged 18 years or older: a randomised, double-blind, placebo-controlled, phase 2 trial. Lancet. 2020; 396(10249):479–488. doi: 10.1016/S0140-6736(20)31605-6.

